# Sensitive quantitative and rapid immunochromatographic diagnosis of clinical samples by scanning electron microscopy - preparing for future outbreaks

**DOI:** 10.1101/2020.05.20.20106864

**Authors:** Hideya Kawasaki, Hiromi Suzuki, Masato Maekawa, Takahiko Hariyama

## Abstract

**Background:** As pathogens such as influenza virus and severe acute respiratory syndrome coronavirus 2 (SARS-CoV-2) can easily cause pandemics, rapid diagnostic tests are crucial for implementing efficient quarantine measures, providing effective treatments to patients, and preventing or containing a pandemic infection. Here, we developed the immunochromatography-NanoSuit^®^ method, an improved immunochromatography method combined with a conventional scanning electron microscope (SEM), which enables observation of immunocomplexes labeled with a colloidal metal.

**Methods and Findings:** The detection ability of our method is comparable to that of real-time reverse transcription-polymerase chain reaction and the detection time is approximately 15 min. Our new immunochromatography-NanoSuit^®^ method suppresses cellulose deformity and makes it possible to easily focus and acquire high-resolution images of gold/platinum labeled immunocomplexes of viruses such as influenza A, without the need for conductive treatment as with conventional SEM. Electron microscopy (EM)-based diagnosis of influenza A exhibited 94% clinical sensitivity (29/31) (95% confidence interval [95%CI]: 78.58–99.21%) and 100% clinical specificity (95%CI: 97.80–100%). EM-based diagnosis was significantly more sensitive (71.2%) than macroscopic diagnosis (14.3%), especially in the lower influenza A-RNA copy number group. The detection ability of our method is comparable to that of real-time reverse transcription-polymerase chain reaction.

**Conclusions:** This simple and highly sensitive quantitative analysis method involving immunochromatography can be utilized to diagnose various infections in humans and livestock, including highly infectious diseases such as COVID-19.

## Introduction

Infections are major threats to humanity. The novel influenza A (H1N1) pdm09 was declared a pandemic in 2009 [1], and the current 2020 severe acute respiratory syndrome coronavirus 2 (SARS-CoV-2) pandemic has had a devastating impact on human health and the world economy. The following factors are crucial for reducing the effects of these infections: 1: delayed invasion of infection into the country because of border measures and quarantine, 2: early containment strategies, 3: early diagnosis enabling appropriate treatment, and 4: broad antibody (IgM, IgG) and/or antigen testing against the virus to assess the spread of the virus. Therefore, rapid diagnostic tests are crucial for disease prevention, treatment, and pandemic containment [2]. Although real-time reverse transcription-polymerase chain reaction (rRT-PCR) is a sensitive method, it is time- consuming, costly, and requires special equipment with professional expertise and high-quality samples. For point-of-care testing, immunochromatography is easier to perform and useful for prompt disease detection, but its sensitivity and specificity are lower than those of rRT-PCR. However, improved specificity has been achieved by using lateral flow biosensors (LFBs) with micro- and nano-materials [3]. Signal readouts based on color, electrochemical signals, magnetic properties, luminescent, and surface-enhanced Raman spectroscopy have been integrated with LFBs for quantification analyses [3,4]. Nevertheless, these nanoparticle sensing methods are indirect. Direct observation of metal nanoparticles by electron microscopy (EM) for clinical use has not been reported because of the complexity of sample preparation and conventional EM operation. We recently reported a method for evaluating multicellular organisms in high vacuum of an EM by encasing them in a thin, vacuum-proof suit, the ‘NanoSuit^®^’ [5], which can impart conductivity to a wet sample to avoid electron charges. Here, we combined the NanoSuit^®^ method with immunochromatography. The new immunochromatography-NanoSuit^®^ method (INSM) suppresses the deformity of the immunochromatography substrate such as cellulose, which causes blurring of particle images, and enables easy focus and acquisition of high-resolution images without the need for additional conductive treatment [6] as with a conventional scanning EM (SEM). In the medical field, using rRT-PCR and INSM as two sensitive rapid diagnostic tests will help maintain patient health. INSM also is a highly sensitive diagnostic tool for several pathogenic infections or other diagnoses.

## Methods

### Ethical statement

The study was approved by the Hamamatsu University School of Medicine ethical committee (No. 19-134), and all methods were performed following relevant guidelines and regulations.

### Immunochromatography kit

The ImunoAce^®^ Flu kit (NP antigen detection), a human influenza commercial diagnosis kit, was purchased from TAUNS Laboratories, Inc. (Shizuoka, Japan). Au/Pt nanoparticles were utilized to visualize the positive lines. A total of 197 clinical samples from patients suspected to be suffering from influenza were provided by a general hospital at the Hamamatsu University School of Medicine for examination using the Flu kit. After macroscopic diagnosis using the Flu kit, the samples were stored in a biosafety box at room temperature (20-25 °C / 68 - 77 °F). The IgM detection immunochromatography kit against SARS-CoV-2 was obtained from Kurabo Industries, Ltd. (Osaka, Japan).

### One step rRT-PCR for influenza A

rRT-PCR for influenza A was performed as described previously using Flu A universal primers [7]. A Ct within 38.0 was considered as positive according to the CDC protocol [8]. The primer/probe set targeted the human RNase P gene and served as an internal control for human nucleic acid as described previously [9].

### SEM image acquisition

The immunochromatography kit was covered with modified NanoSuit^®^ solution based on previously published components [5] (Nisshin EM Co., Ltd., Tokyo, Japan), placed first onto the wide stage of the specimen holder, and then placed in an Lv-SEM (TM4000Plus, Hitachi High-Technologies, Tokyo, Japan). Images were acquired using backscattered electron detectors with 10 or 15 kV at 30 Pa.

### Particle counting

In fields containing fewer than 50 particles/field, the particles were counted manually. Otherwise, ImageJ/Fiji software was used for counting. ImageJ/Fiji uses comprehensive particle analysis algorithms that effectively count various particles. Images were then processed and counting was performed according to the protocol [10].

### Diagnosis and statistics

The EM diagnosis and criteria for a positive test were defined as follows: particle numbers from 6 fields from the background area and test-line were statistically analyzed using the *t*-test. If there were more than 5 particles in one visual field and a significant difference (P < 0.01) was indicated by the *t*-test, the result was considered as positive. Statistical analysis using the *t*-test was performed in Excel software. Statistical analysis of the assay sensitivity and specificity with a 95% confidence interval (95% CI) was performed using the MedCalc statistical website. The approximate line, correlation coefficient, and null hypothesis were calculated with Excel software.

## Results

To investigate the sensitivity and specificity of immunochromatography using the NanoSuit^®^ method, an influenza diagnostic kit (TAUNS Laboratories, Inc.) was prepared (**Fig. 1*A***). Two specific antibodies were used: one (anti-mouse IgG or anti-influenza A NP) was immobilized on chromatographic paper, whereas the other was labeled with colloidal gold/platinum (100−200 nm in diameter) and infiltrated into the sample pad. The kit was completed by attaching the sample pad at the end of the membrane. When the clinical sample in lysis buffer (150 DL) was placed on the sample pad, the virus antigen in the sample formed an immunocomplex with the colloidal gold/platinum- labeled antibody, which subsequently formed an immune complex with the antibody immobilized on the membrane, resulting in the generation of colored lines and indicating the presence of the antigen of interest in the sample (**Fig. 1*B* top, middle**). After the reaction, NanoSuit^®^ solution (100 μL) was added upstream of the test-line (**Fig. 1*B* middle**), forming a thin NanoSuit^®^ liquid layer (pale blue) (**Fig. 1*B* bottom**). The kit was positioned on the sample stage in the sample chamber of the SEM as close and parallel as possible to the camera (**Fig. 1*C***). The observation position of the test- line and background area were determined at fixed distances from the control-line border. The number of Au/Pt particles at the test-line and background area were counted in 6 fields of view at ×1200 (**Fig. 1*D***). Without NanoSuit^®^ treatment, swelling of the cellulose and residual liquid present due to electron beam energy were observed (**Fig. 2*A***). In contrast, following NanoSuit^®^ treatment, the cellulose membrane showed little or no swelling (**Fig. *2B***).

**Figure 1.**
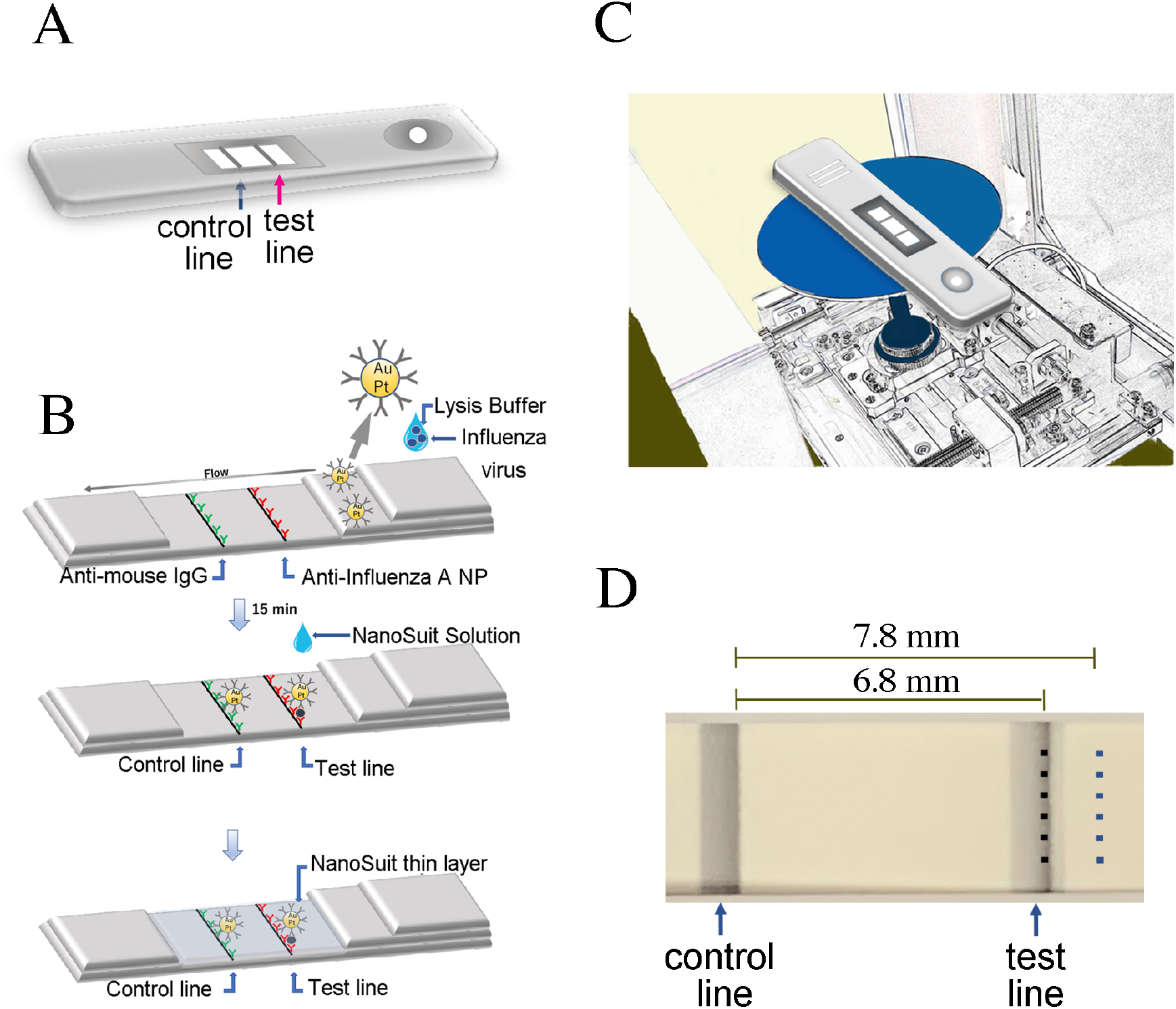
Sensitivity and specificity of immunochromatography using the NanoSuit^®^ method. (***A***) Kit representation showing test and control lines. (***B***) Schematic diagram of the gold/platinum (Au/Pt)-Ab conjugate-linked rapid immunochromatographic kit. The immune-complex reacted with the anti-influenza A nucleoprotein (NP) at the test line and anti-mouse IgG at the control-line (top, middle). A NanoSuit^®^ thin layer was formed after NanoSuit^®^ treatment (bottom). (***C***) Kit placement in the SEM chamber. (***D***) Determination of the observation positions. Six fields were randomly selected in the test line and background areas.

**Figure 2.**
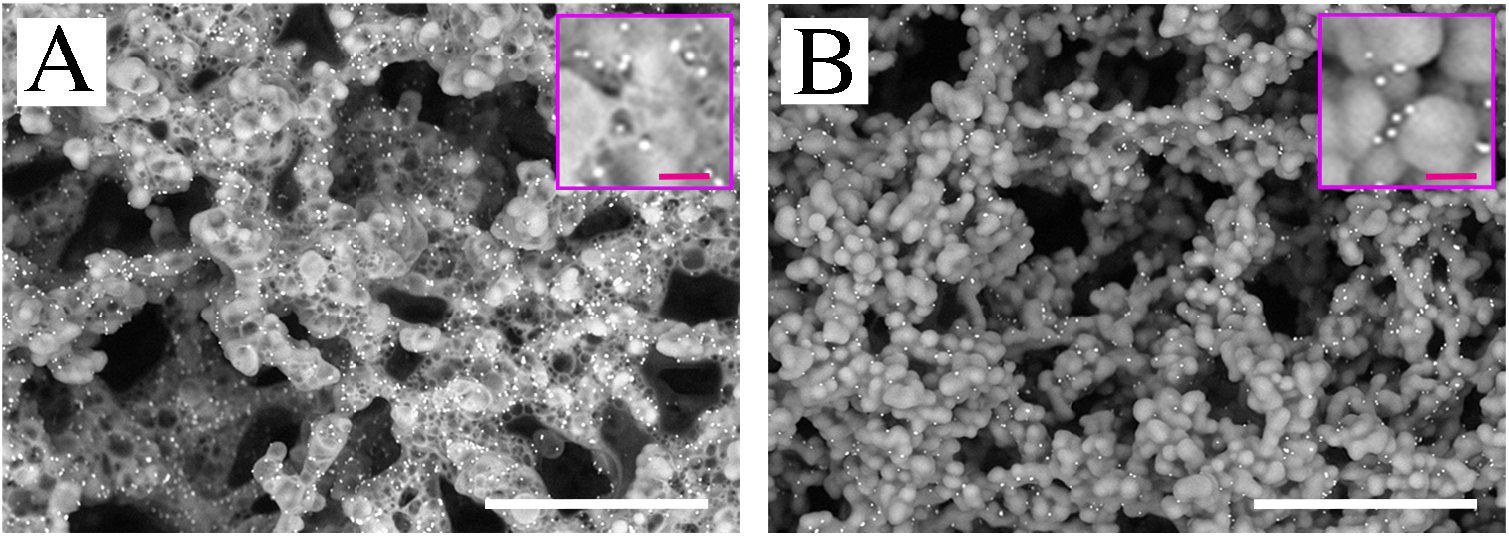
(***A, B***) Images of cellulose and Au/Pt-labeled immunocomplex with immobilized antibody without (***A***) and with NanoSuit treatment (***B***). Scale bars in (***A***) and (***B***) = 15 μm. Inset is magnified image. Scale bars 600 nm.

The SEM images of the test lines and background were compared. **Figure 3*A, D*** show the “macroscopic diagnosis-positive” test-line. **Figure 3*B, E*** are images of the “macroscopic diagnosis-negative” and “EM diagnosis-positive” test-line. **Figure 3*C, F*** are the images of the background. Au/Pt particles (arrows) of the test-line were clearly visualized (**Fig. 3*A, B, D, E***) compared to background areas of the cellulose membrane (**Fig. 3*C, F***). Au/Pt particle counting was performed by using ImageJ/Fiji software for macroscopic diagnosis-positive samples. Manual counting was performed for the images of “background” and “macroscopic diagnosis negative and SEM diagnosis-positive” samples as well as “EM-negative” samples. Another immunochromatography diagnosis kit for detecting IgM antibodies against SARS-CoV-2 from Kurabo Industries, Ltd. was tested using the NanoSuit^®^ method (**S1*A* Fig**). The cellulose membrane image without the immunocomplex was clearly visualized after NanoSuit^®^ treatment (**S1*B* Fig**). Approximately 25-nm countable gold nanoparticles (GNPs) of the control-line were detected (**S1*C* Fig)**.

**Figure 3.**
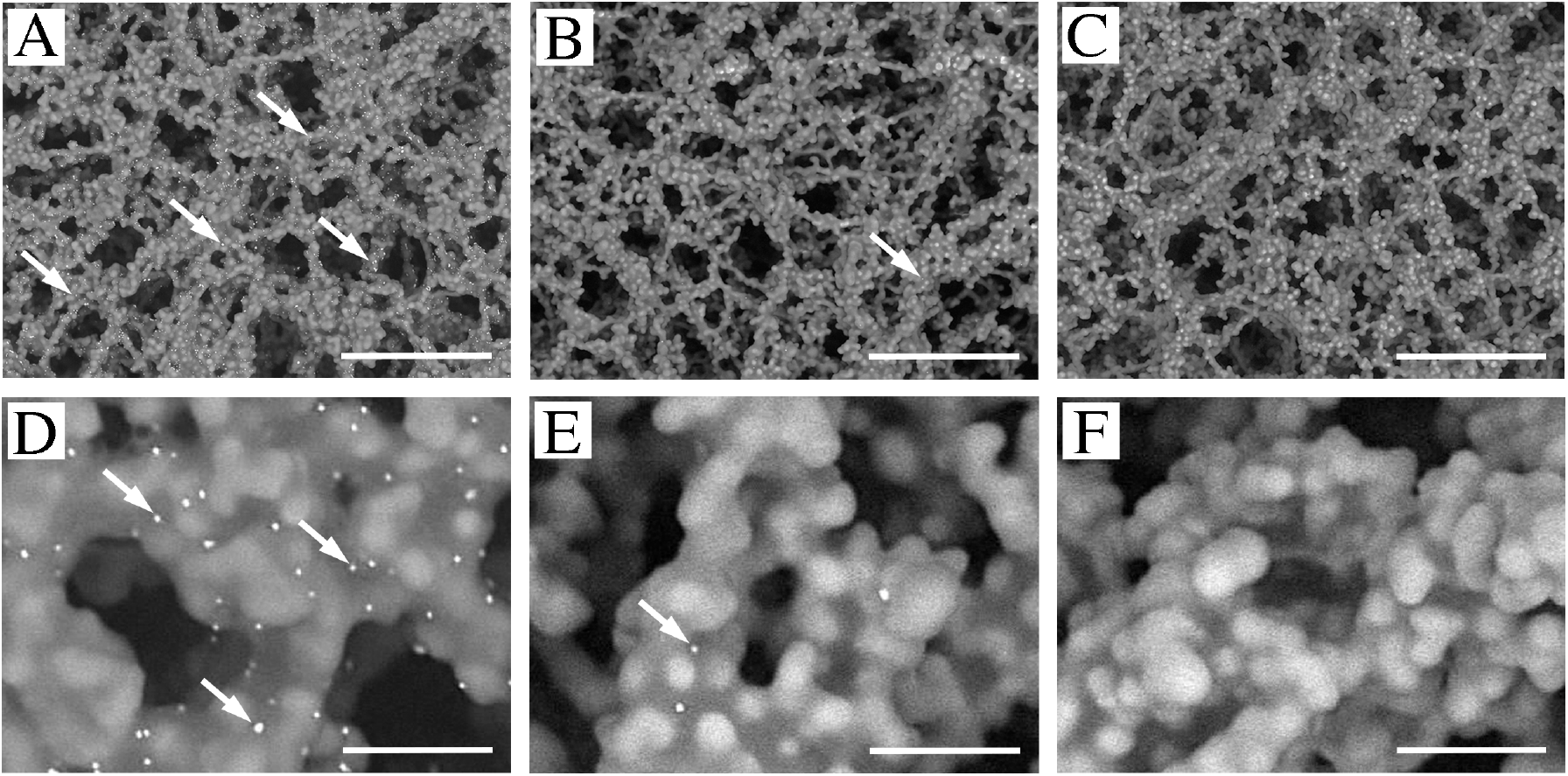
Comparison of SEM images of test lines and of background. **(*A, D***) images of “macroscopic diagnosis-positive” test-line. **(*B, E)*** Images of “macroscopic diagnosis-negative” and “EM diagnosis-positive” test-line. **(*C, F*)** Images of “background”. White arrows indicate representative Au/Pt particles. Scale bars in (***A, B, C***) = 30 μm and (***D, E, F***) = 3 μm.

Diagnoses based on macroscopic, EM, and rRT-PCR results were compared using the 197 influenza-suspected clinical samples. rRT-PCR for influenza A was performed with the same clinical pharyngeal swab samples as used in immunochromatography. To examine the relationship between the influenza copy number and rRT-PCR threshold, serial dilutions were prepared (**S2*A* Fig**). A calibration curve (**S2*B* Fig**) was drawn to determine the relationship between the copy number and cycle threshold (Ct) (**S2*C* Fig**). In our assay system, Ct ≤ 38.0 was calculated to be ≥ 151.4 copies/reaction.

The quantitative relationship between particle counts/fields (log10) and Ct are shown as a scatter diagram. The correlation coefficient of Ct and particle counts/field was -0.803, which was significant (p = 3.79E-08) and the null hypothesis was rejected (**Fig. 4**) (**Supplemental data**).

**Figure 4.**
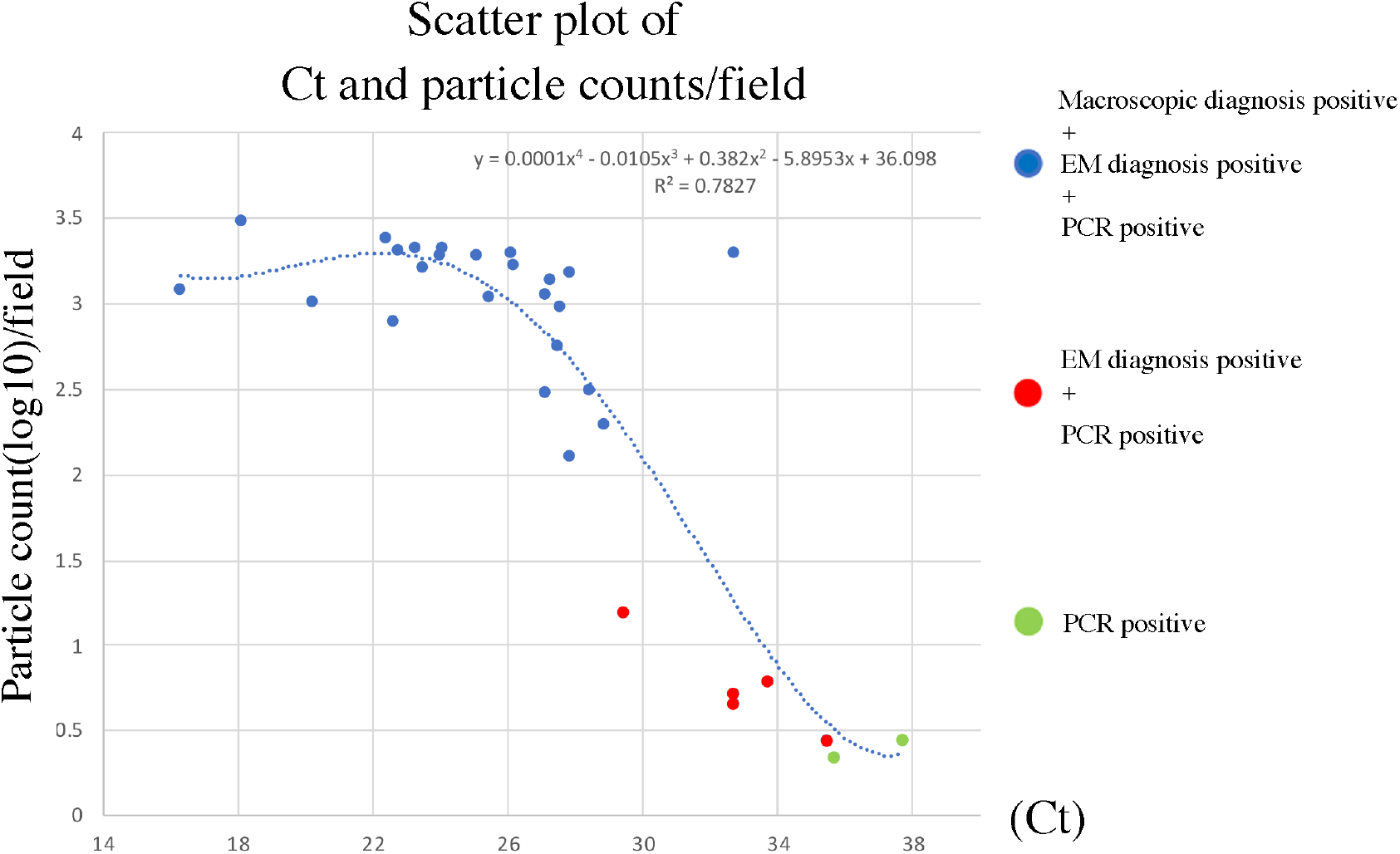
Scatter plot of Ct and particle counts/field. Blue dot represents triple-positive (“macroscopic diagnosis-positive” and “EM diagnosis-positive” and “PCR diagnosis-positive”). Red dot represents double-positive (“EM diagnosis-positive” and “PCR diagnosis-positive”). Green dot represents single-positive (“PCR diagnosis-positive”). Blue dot line is approximate curve. Vertical axis: particle counts (log10)/field. Horizontal axis: Ct of influenza A.

The EM diagnosis for influenza A showed 94% clinical sensitivity (29/31) (95% confidence interval [95%CI]: 78.58–99.21%) and 100% clinical specificity (95%CI: 97.80–100%) (**Table 1**) (**Supplemental data**), as well as a strong correlation (kappa; 0.99) compared with the results obtained by rRT-PCR (14.0 ≤ Ct ≤ 38.0) (**Table 2**) (**Supplemental data**). In contrast, standard macroscopic diagnosis showed 77% clinical sensitivity (24/31) (95%CI: 58.90–90.41%) and 100% clinical specificity (95%CI: 97.80–100%) (**Table 1**) (**Supplemental data**), along with a strong correlation (kappa; 0.96) compared with the results obtained by rRT-PCR (14.0 ≤ Ct ≤ 38.0) (**Table 2**)(**Supplemental data**).

**Table 1.** Clinical diagnostic performance of EM assay. Thirty-one influenza A-rRT-PCR-positive samples were used to determine the sensitivity of the assays, and 166 influenza rRT-PCR-negative samples were used to determine the specificity of the assays.

| Influenza A<br>rRT-PCR | Ct | Macroscopic diagnosis |  | Electron microscopic diagnosis |  |
| --- | --- | --- | --- | --- | --- |
|  |  | Sensitivity | Specificity | Sensitivity | Specificity |
| Positive<br>(n=31) | 14.0≤Ct<22.0 | 100% (3/3) |  | 100% (3/3) |  |
|  | 22.0≤Ct<30.0 | 95.2% (20/21) |  | 100% (21/21) |  |
|  | 30.0≤Ct≤38.0 | 14.3% (1/7) |  | 71.4% (5/7) |  |
|  | 14.0≤Ct≤38.0 | 77.4% (24/31) |  | 93.5% (29/31) |  |
| Negative<br>(n=166) | Ct>38.0 |  | 100% (166/166) |  | 100% (166/166) |

**Table 2.**
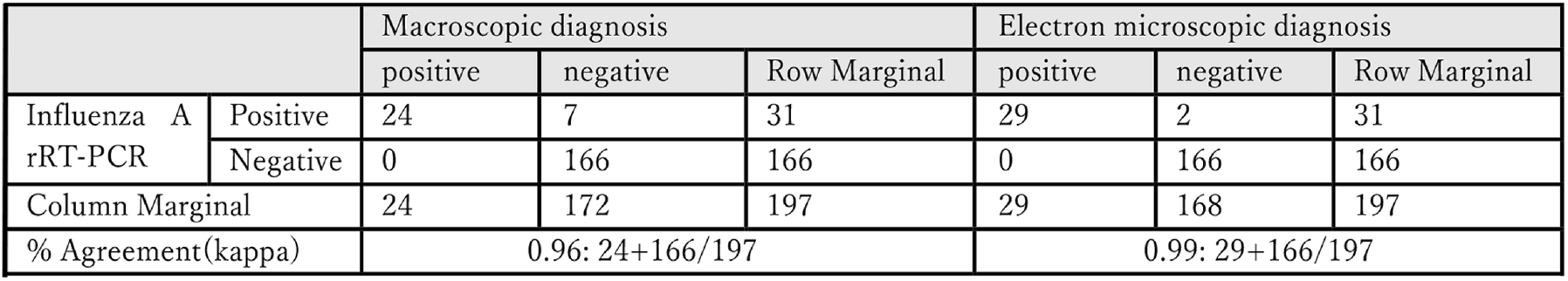
Comparison of EM diagnosis with rRT-PCR and macroscopic diagnosis by immunochromatography.

|  |  | Macroscopic diagnosis |  |  | Electron microscopic diagnosis |  |  |
| --- | --- | --- | --- | --- | --- | --- | --- |
|  |  | positive | negative | Row Marginal | positive | negative | Row Marginal |
| Influenza A<br>rRT-PCR | Positive | 24 | 7 | 31 | 29 | 2 | 31 |
|  | Negative | 0 | 166 | 166 | 0 | 166 | 166 |
| Column Marginal |  | 24 | 172 | 197 | 29 | 168 | 197 |
| % Agreement(kappa) |  | 0.96: 24+166/197 |  |  | 0.99: 29+166/197 |  |  |

## Discussion

Rapid diagnostic tests show variable assay performance with sensitivities of 10– 70% and up to 90% specificity compared to standard rRT-PCR-based assays [11]. Their sensitivity has been improved by employing europium nanoparticles, which show 82.59% sensitivity and 100% specificity for clinically evaluated influenza A (H1N1) [12]. The use of silver amplification immunochromatography in influenza virus detection kits showed 91.2% sensitivity and 95.8% specificity [13]. Moreover, the rapid fluorescent immunochromatographic test employing CdSe/CdS/ZnS quantum dots showed 93.75% clinical sensitivity and 100% clinical specificity [14]. However, the potential toxic effects of cadmium-based quantum dots are controversial [15]. INSM is safe and showed the highest sensitivities (94% clinical sensitivity, 100% clinical specificity) in this study. Generally, overall sensitivity depends on the distribution of each sample’s pathogen-copy number in the investigated group. Our study revealed that EM diagnosis was significantly more sensitive (71.2%) than macroscopic diagnosis (14.3%) in the lower copy number group (30.0 ≤ Ct ≤ 38). The detection ability of our method is comparable to that of rRT-PCR (**Fig. 4**). Theoretically and practically, our method shows the highest detection performance as an immunochromatographic diagnostic method. More sensitive immunochromatographic products may be developed in future through further improvements such as by optimizing the antigen concentration.

PCR has recently been used as a main diagnostic tool for SARS-CoV-2. The introduction of immunochromatography analysis using the SARS-CoV-2 antigen has greatly changed testing practices. All immunochromatographic-negative SARS-CoV-2-suspected samples are recommended for analysis by PCR. Highly sensitive immunochromatographic tests for infectious diseases can greatly reduce the number of PCR samples to be analyzed. Furthermore, Cohen and Kessel reported high false-positive rates of RT-PCR testing for SARS-CoV-2 using clinical samples. Overall, 336 of 10,538 negative samples (3.2%) were reported as positive. In contrast, <0.6–7.0% false-positive rates in external quality assessments of RNA virus assays were reported for influenza A. The amplification of nucleic acids makes PCR-based assays highly sensitive but highly vulnerable to minute levels of sample contamination which can produce false-positive results indistinguishable from true-positive results [16]. The same pitfall may be found in Loop-Mediated Isothermal Amplification (LAMP) method. We propose that macroscopic immunochromatographic-negative and PCR- or LAMP-positive cases should be compared with highly sensitive immunochromatographic data such as that obtained by INSM as well as clinical data to reduce false-positive cases.

Although ImageJ/Fiji is useful software, it should be further developed to increase its reliability. The development of an automated GNP counting system to replace manual counting using deep learning is being evaluated by our team. Furthermore, clinical application of using an automated inexpensive SEM for immunochromatography in combination with INSM for SEM shows potential. Among the commonly used micro/nano-particles in immunochromatography, colloidal GNP is the most widely used [3]. GNP is safe and can be easily conjugated with biomolecules that retain their biochemical activity upon binding. Therefore, developing new GNP-based LFBs may be easier and faster than developing micro/nano-materials and can be adapted quickly for new emerging infections such as SARS-CoV-2 [17]. Investigation of INSM using smaller GNPs is currently underway. INSM is applicable to all LFBs, particularly in emergency tests to evaluate troponin, brain natriuretic peptide, and procalcitonin as measured by immunochromatography.

Diagnosis using the INSM shows high sensitivity, as it allows for direct particle observation. Our results indicate that INSM can be used for automated quantitative measurement in any immunochromatographic tests including recently developed SARS-CoV-2 antigen or IgM/IgG antibodies tests against SARS-CoV-2. This method can be used to promptly diagnose new emerging infections including those in livestock and promote innovations in assays using LFBs.

## Data Availability

The authors confirm that the data supporting the findings of this study are available within the article

## Acknowledgments

The authors thank Noriko Aoki (TAUNS Laboratories, Inc.) for providing rRT-PCR data on influenza A and Takafumi Miwa and Takumi Tandou (Hitachi, Ltd. Research & Development Group Nano-process Research Department) for advising on SEM. We also thank the clinical laboratory center of Hamamatsu University School of Medicine, University Hospital for providing influenza immunochromatography test strips after routine examinations. This work was supported by JST START (grant number 714 [to H.K.]), JSPS KAKENHI (grant numbers JP17K08784 [to H.K.] and JP18H01869 [to T.H.]), and AMED (grant number A508 [to H.K.]).

## Competing interests

The authors declare no competing interests.

### Abbreviations

EM: Electron microscopy
LAMP: loop-mediated isothermal amplification
SEM: scanning electron microscope
GNP: gold nanoparticles
INSM: immunochromatography-NanoSuit^®^ method
PCR: polymerase chain reaction
rRT-PCR: real-time reverse transcription-polymerase chain reaction
IgG: immunoglobulin G
IgM: immunoglobulin M

## Supplemental Figures and Data

**Supplementary Figure 1.**
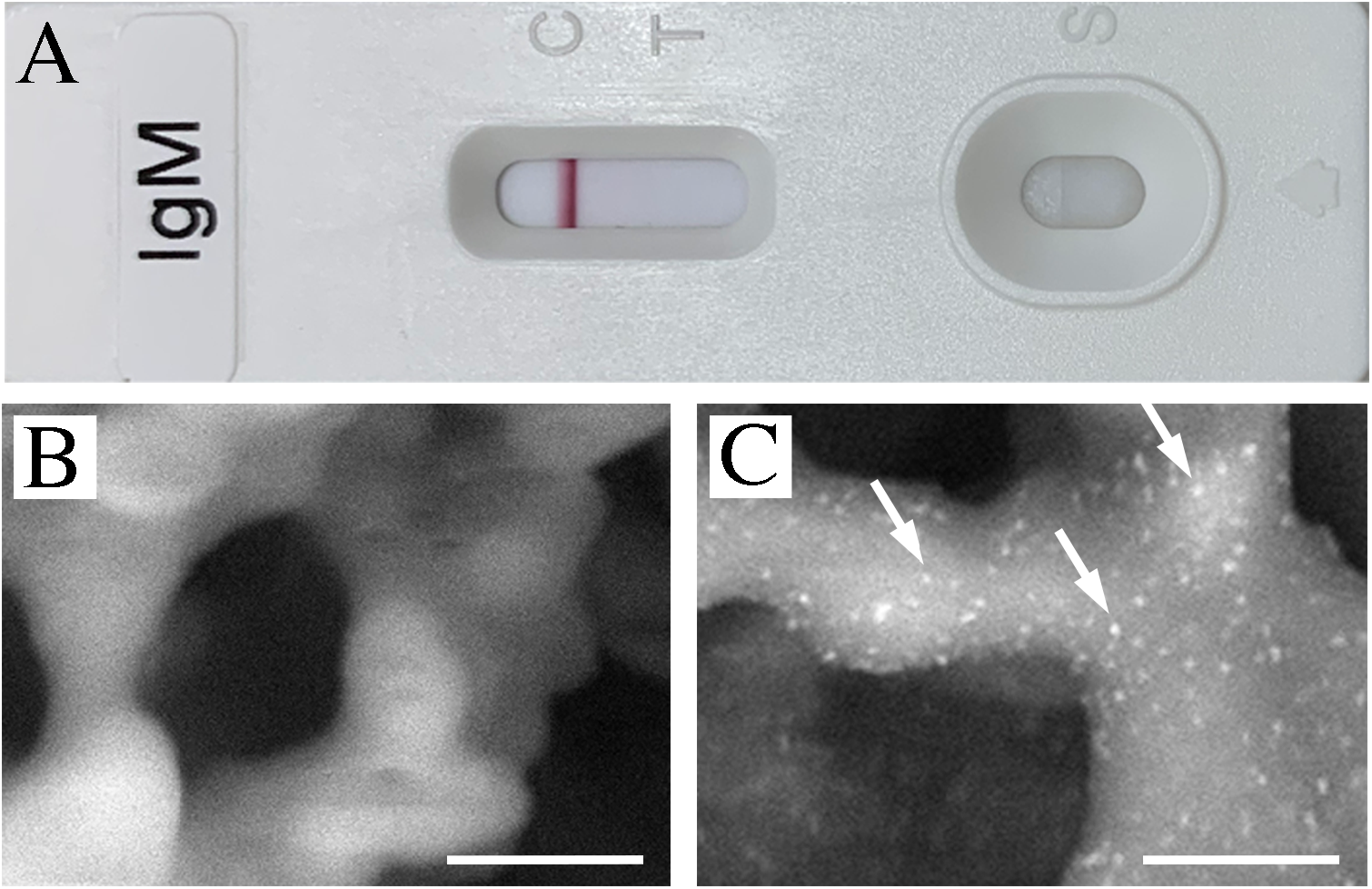
(***A***) Immunochromatography diagnosis kit for IgM antibody against SARS-CoV-2. (***B***) Background image. (***C***) Multiple gold nanoparticles (GNPs) (approximately 25-nm diameter) of the control-line on cellulose. White arrows indicate representative GNPs. Scale bars in (***B***) and (***C***) = 600 nm.

**Supplementary Figure 2.**
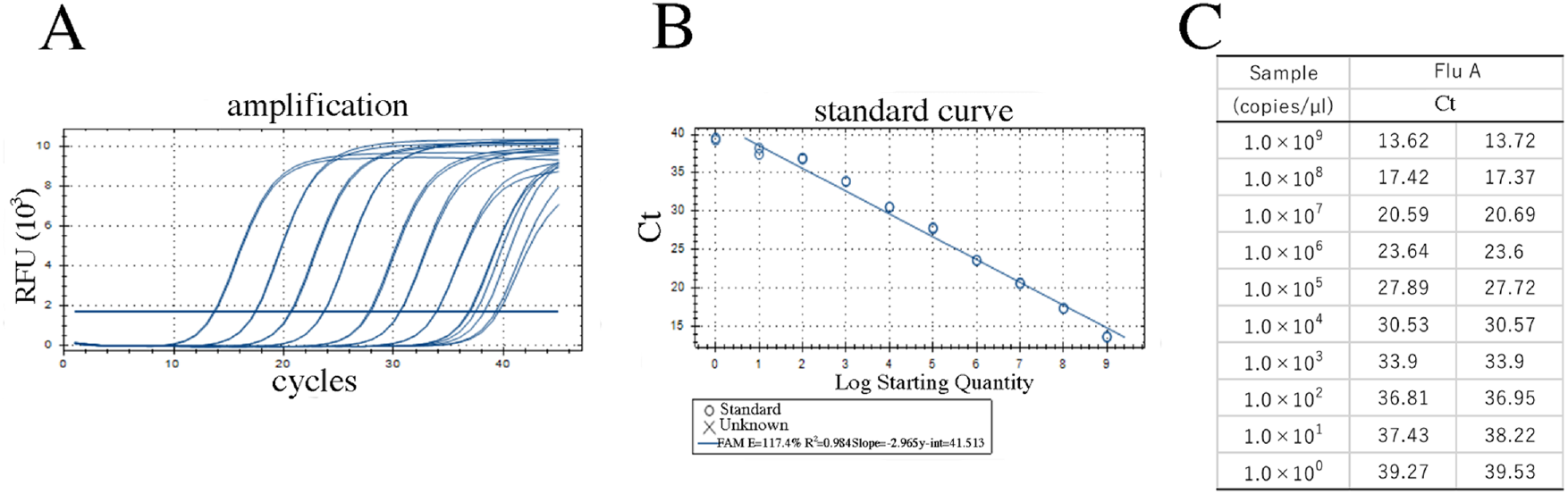
Comparison of macroscopic, electron microscopic (EM) and rRT-PCR influenza A diagnoses. (***A***) rRT-PCR amplification curve for influenza A. (***B***) Standard curve for rRT-PCR of influenza A. (***C***) Relationship between cycle threshold (Ct) and sample copy numbers/reaction.

## Supplemental Data

Comparison of among macroscopic diagnosis, EM diagnosis and rRT-PCR diagnosis. BG: Background, TL: Test Line

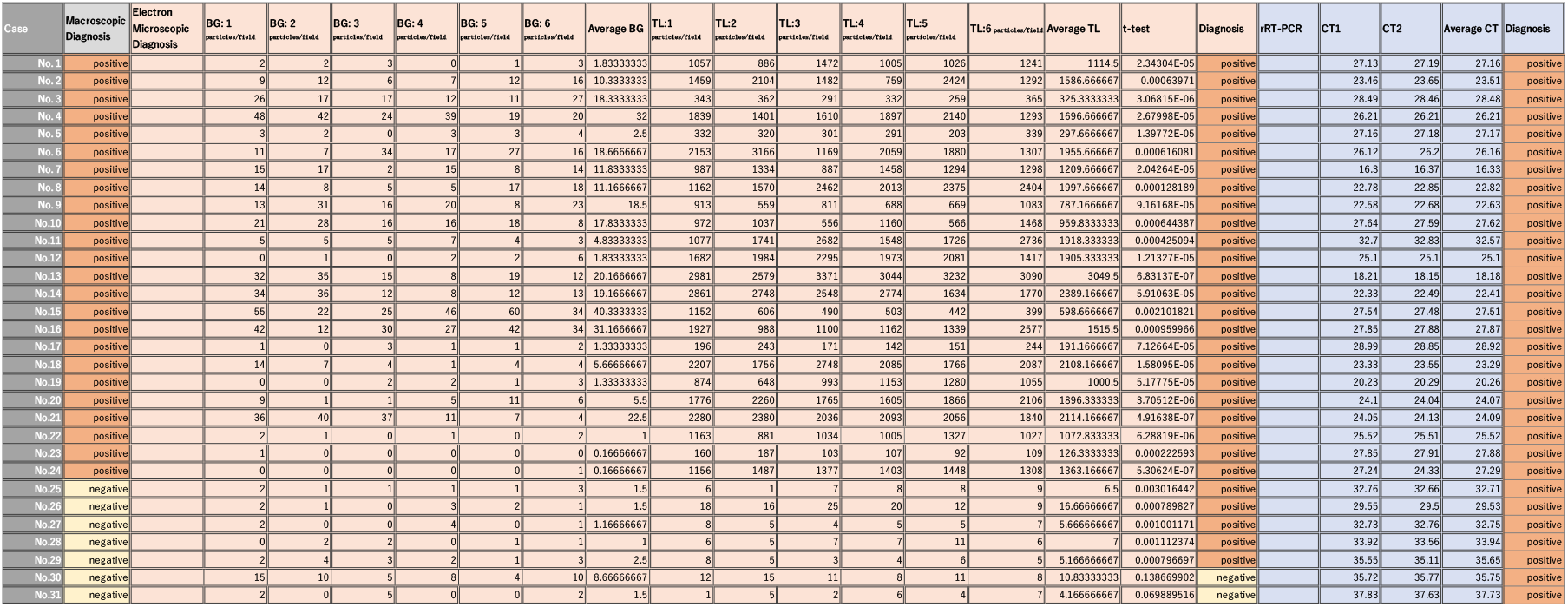

## Notes

### Competing Interest Statement

The authors have declared no competing interest.

### Funding Statement

This work was supported by JST START (grant number 714 [to H.K.]), by JSPS KAKENHI (grant numbers JP17K08784 [to H.K.] and JP18H01869 [to T.H.]), and by AMED (grant number A508 [to H.K.]).

## References

[1] Swerdlow DL, Finelli L. Preparation for possible sustained transmission of 2019 novel coronavirus: lessons from previous epidemics. JAMA. 2020;323: 1129–1130.

[2] Ravina R, Dalal A, Mohan H, Prasad M, Pundir CS. Detection methods for influenza A H1N1 virus with special reference to biosensors: a review. Biosci Rep. 2020;40.

[3] Huang Y, Xu T, Wang W, Wen Y, Li K, Qian L, et al. Lateral flow biosensors based on the use of micro- and nanomaterials: a review on recent developments. Mikrochim Acta. 2019;187: 70.

[4] Urusov AE, Zherdev AV, Dzantiev BB. Towards lateral flow quantitative assays: detection approaches. Biosensors (Basel). 2019;9.

[5] Takaku Y, Suzuki H, Ohta I, Ishii D, Muranaka Y, Shimomura M, et al. A thin polymer membrane, nano-suit, enhancing survival across the continuum between air and high vacuum. Proc Natl Acad Sci USA. 2013;110: 7631–7635.

[6] Kawasaki H, Itoh T, Takaku Y, Suzuki H, Kosugi I, Meguro S, et al. The NanoSuit method: a novel histological approach for examining paraffin sections in a nondestructive manner by correlative light and electron microscopy. Lab Invest 2020;100: 161–173.

[7] de-Paris F, Beck C, Machado ABMP, Paiva RM, da Silva Menezes D, de Souza Nunes L, et al. Optimization of one-step duplex real-time RT-PCR for detection of influenza and respiratory syncytial virus in nasopharyngeal aspirates. J Virol Methods. 2012;186: 189–192.

[8] CDC. Division CDC Human Influenza Virus Real-Time RT-PCR Diagnostic Panel (CDC Flu rRT-PCR Dx Panel). 2014. Available from: https://journals.plos.org/plosone/article/file?type=supplementary&id=info:doi/10.1371/journal.pone.0201248.s007.

[9] WHO Collaborating Centre for Influenza, GA. CDC protocol of realtime RTPCR for influenza A (H1N1) 2009. Available from: https://www.who.int/csr/resources/publications/swineflu/realtimeptpcr/en/.

[10] O’Brien J, Hayder H, Peng C. Automated quantification and analysis of cell counting procedures using ImageJ plugins. J Vis Exp. 2016;17: 54719.

[11] Vemula SV, Zhao J, Liu J, Wang X, Biswas S, Hewlett I. Current approaches for diagnosis of influenza virus infections in humans. Viruses. 2016;8: 96.

[12] Yu ST, Bui CT, Kim DTH, Ngyuen AVT, Trinh TTT, Yeo SJ. Clinical evaluation of rapid fluorescent diagnostic immunochromatographic test for influenza A virus (H1N1). Sci Rep. 2018;8: 13468.

[13] Mitamura K, Zhimizu H, Yamazaki M, Ichikawa M, Nagai K, Katada J, et al. Clinical evaluation of highly sensitive silver amplification immunochromatography systems for rapid diagnosis of influenza. J Virol Methods. 2013;194: 123–128.

[14] Nguyen AVT, Dao TD, Trinh TTT, Choi DY, Yu ST, Park H, et al. Sensitive detection of influenza a virus based on a CdSe/CdS/ZnS quantum dot-linked rapid fluorescent immunochromatographic test. Biosens Bioelectron. 2020;155: 112090.

[15] Oh E, Liu R, Nel A, Gemill KB, Bilal M, Cohen Y, et al. Meta-analysis of cellular toxicity for cadmium-containing quantum dots. Nat Nanotechnol. 2016;11: 479–486.

[16] Cohen AN, Kessel B. False positives in reverse transcription PCR testing for SARS-CoV-2. medRxiv. 2020.

[17] Sheridan C. Fast, portable tests come online to curb coronavirus pandemic. Nat Biotechnol. 2020;38: 515–518.

